# Relationships of visual impairment and eye conditions with imaging markers, cognition, and diagnoses of dementia: a bi-directional Mendelian randomization study

**DOI:** 10.1101/2024.01.05.24300912

**Authors:** Erin L Ferguson, Mary Thoma, Peter Buto, Jingxuan Wang, M. Maria Glymour, Thomas J Hoffmann, Hélène Choquet, Shea J Andrews, Kristine Yaffe, Kaitlin Casaletto, Willa D Brenowitz

**Author notes:** **Corresponding Author**: Erin Ferguson.

## Abstract

**Objective:** To evaluate the causal relationships between visual acuity, eye conditions (focusing on cataracts and myopia), and Alzheimer disease (AD) and related dementias.

**Design:** Cohort and two sample bi-directional mendelian randomization (MR) study.

**Setting:** UK Biobank participants and summary statistics from previously published genome-wide association studies on cataract, myopia, and AD.

**Participants:** UK Biobank participants (n=304,953) aged 55-70 without dementia at baseline, underwent genotyping, reported on eye conditions, and a subset completed visual acuity exams (n=113,756) or brain imaging (n=36,855)

**Main outcome measures:** All-cause dementia, AD, and vascular dementia (VaD) identified from electronic medical records.

**Results:** The sample averaged 62.1 years (SD=4.1) of age at baseline, 4.7% had cataracts, and 3.9% had worse than 20/40 vision. History of cataracts (HR=1.18, 95% CI: 1.07 to 1.29) and 20/40 vision (HR=1.35, 95% CI: 1.06 to 1.70) were associated with higher hazard of all-cause dementia. In MR analyses to estimate causal effects, cataracts increased risk of VaD inverse-variance weighted (OR=1.92, 95% CI: 1.26-2.92) borderline increased all-cause dementia (OR =1.21, 95% CI: 0.98 to 1.50) but not AD (OR=1.01, 95% CI: 0.97-1.06). There was no significant association between observed or genetic risk for myopia and dementia. In MR for reverse causality using genetic risk for AD, AD was not significantly associated with cataracts (inverse-variance weighted OR=0.99, 95% CI: 0.96 to 1.01). Genetic risk for cataracts were associated with smaller total brain (β= −597.4 mm^3^, 95% CI: −1077.9 to −117.0) and grey matter volumes (β= −375.2 mm^3^, 95% CI: −680.1 to −70.2), but not other brain regions or cognition.

**Conclusions:** Our findings suggest cataracts increase risk of dementia and may reduce brain volume. This lends further support to the hypothesis that cataract extraction may reduce risk for dementia.

## INTRODUCTION

Visual impairment and ocular disease are emerging as a potential modifiable risk factors for Alzheimer disease (AD) and related dementias (ADRD).(1) Vision impairment, including self-reported impairment,(2,3) due to the presence of eye conditions like cataracts,(4,5) and poor visual acuity,(6–8) are associated with increased risk of dementia or cognitive decline with age. If causal, this relationship would be clinically meaningful, as visual impairment could be reversed through clinical intervention. For example, observational studies have reported that cataract surgery is associated with decreased risk of dementia.(4,9)

However, establishing the causality of this relationship is methodologically difficult. Both vision impairment and ADRD can develop slowly over the life course without distinct clinical onsets, they have many shared risk factors such as age and cardiovascular risk factors,(10,11) and early ADRD could affect visual processing (reverse causation).(12) Additionally, previous studies, especially those using electronic health records (EHR), are often subject to unmeasured confounding by important social factors relevant for dementia research, like education and income,(13–15) or confounding by indication.(16) Given the uncertainty of the direction of this relationship, alternative approaches are needed to evaluate this question and to inform whether vision care could help delay onset of ADRD.

Methods using genetic variation to evaluate the link between visual impairment and ADRD risk have the potential to address this gap and have not been used extensively. Genetic variants for eye conditions and for AD,(17–19) which are well established, can be leveraged to evaluate causal relationships, because genetic variants are likely independent of key confounders of the relationships of interest. Mendelian randomization (MR) uses these genetic variants, single nucleotide polymorphisms (SNPs), as instruments, allowing for an unbiased estimate of the causal effects provided certain assumptions are met.(20,21) However, few studies have used MR methods to evaluate relationships between vision impairments and dementia. One prior study reported null findings for the effect of cataracts on AD, however, this study did not examine vision impairments or dementia more broadly.(22) Furthermore, previous studies have not investigated related indicators of ADRD, such as neuroimaging markers, which would provide convergent evidence for a causal relationship and could help elucidate any underlying biologic pathways.

We evaluated the relationships between visual impairment, cataracts, and ADRD outcomes, using data from the UK Biobank. With recent interest in visual acuity(23) and cataracts(9) as risk factors for dementia, along with sample size limitations of other visual conditions, we focused our genetic analyses on cataracts and myopia. Using MR, we first used genetic risk for AD to evaluate whether shared genetics or incipient AD increases the risk of later vision impairment or eye conditions (**fig 1A**). Second, we used genetic risk for eye conditions (cataracts and myopia) to evaluate whether incipient eye conditions causally increase the risk of AD and related dementias, vascular dementia (VaD), or AD (**fig 1B**). Finally, to inform us on biological pathways, we evaluated associations between eye conditions and brain magnetic resonance imaging (MRI) outcomes.

**Fig 1.**
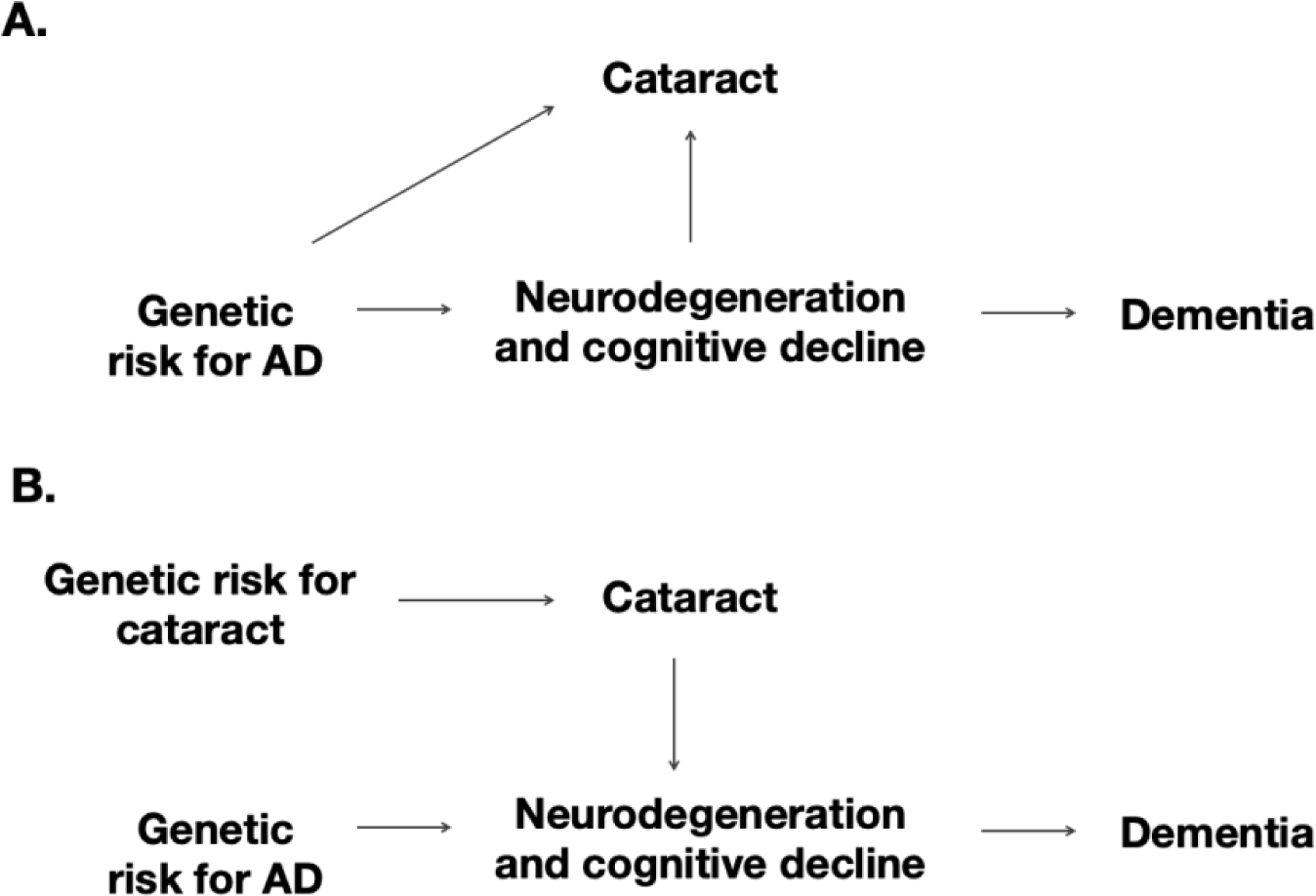
Directed acyclic graph of two hypothesized relationships between cataracts (or other eye conditions) and brain regions, leading to dementia. Using Mendelian randomization, we can use variations in genetic risk to distinguish between two causal scenarios. If incipient neurodegeneration or other physiological markers cause cataracts to form, then genetic risk for AD should be associated with later cataracts (Panel A). If cataracts has independent effects on processes leading to dementia (i.e. cataracts are a causal risk factor for dementia), then genetic risk for AD should be independent of cataracts and that genetic risk for cataracts would be associated with dementia (Panel B).

## METHODS

### Study population

The UK Biobank is an ongoing cohort study that has enrolled over 500,000 individuals aged 40-69 years from 2006 to 2010 across the United Kingdom.(24) Cohort members provided biological samples and detailed survey responses, with subsets completing structural brain MRI, visual acuity exams, and cognitive assessments. Visual acuity exams and cognitive assessments were collected at baseline visits, brain imaging occurred at a later visit, and we had longitudinal follow-up for dementia from electronic health records through November 2021.

We included UK Biobank participants 55 years or older (n=308,272) to ensure this population was susceptible to cataracts and cognitive change or decline over follow-up. We excluded participants with dementia at baseline (n=192, [0.06%]) or without self-reported visual health history data (n=3,127, [1.0%]) for an analytic sample of 304,953. Analytic subcohorts with complete covariate data were created for respective analyses due to data availability. For analyses using neuroimaging outcomes, we restricted to participants with available brain MRI data (n=36,591–36,855). Analyses using visual acuity or myopia exposure were restricted to individuals who completed visual acuity exams at their initial assessment visit (n=69,852-71,429). Analyses with cognitive score outcomes were restricted to individuals with completed pairs matching and reaction time assessments (n=301,362-302,402).(25) Analyses using genetic risk scores or Mendelian randomization were restricted to those with European genetic ancestry and those 60 years and older (n=183,760, 84.4% of analytic sample).

### Eye disorders and visual acuity

Participants self-reported their history of eye problems (including cataracts, glaucoma, age-related macular degeneration (AMD), and diabetic retinopathy) during their baseline visit. In the same baseline survey, participants self-reported their history of cataract surgery. For analyses in which cataracts were the outcome, we used additional diagnoses of senile cataracts that were obtained from linked EHR data through the end of 2021. In the subset completing eye and vision exams (n=113,756), presence of myopia was determined from refraction error, measured by non-cycloplegic autorefraction.(26,27) In the same ophthalmic assessment battery, visual acuity was measured as the logarithm of the minimum angle of resolution (logMAR) chart.(27) This exam was completed for both eyes and the logMAR measure from the better eye was used. Binary 20/40 visual acuity was calculated using a logMAR cutoff of 0.3. Due to recent literature and interest in cataract and visual acuity, and also because of the small sample size of other visual conditions in UKB, our primary exposures of interest were cataracts and myopia.

### Dementia and cognition

Incident dementia cases, including Alzheimer’s disease, vascular dementia, and non-specific dementia, were ascertained and defined two ways to improve the positive predictive value of diagnoses.(28) First, we used the UK Biobank derived algorithmically-defined outcome for all-cause dementia.(29) Second, we extracted dementia diagnoses from linked primary care and in-patient EHR through 2021 using ICD codes (**supplemental table 1**). In sensitivity analyses, we restricted the outcome to dementia subtypes, including AD or VaD.

We also evaluated whether cataracts and myopia were associated with preclinical changes in cognition. Cognitive assessments were conducted in person at the participant’s initial assessment visit. We used two available measures of cognition from this initial visit: 1) average reaction time from the ‘Snap’ test and 2) number of incorrect matches from the Pairs matching task averaged over two rounds.(30) These assessments measure reaction time and visual memory, and were selected because 1) they had a larger sample size than other cognitive phenotypes, and 2) because genetic risk of AD has previously been shown to be associated with these phenotypes.(31)

### Brain Magnetic Resonance Imaging Volumes

To assess whether eye conditions were associated with markers of dementia not encapsulated in a diagnosis or cognitive performances, we selected five neuroimaging regions of interest as secondary outcomes. Based on previous research showing their association with cognitive decline, we included measures of total brain volume,(32) total grey matter volume,(33,34) hippocampal volume (left and right hemispheres),(33,35) white matter hyperintensity volume,(36) and mean cortical thickness in an AD signature region.(37) The AD signature region is a surface area-weighted average of cortical thicknesses across six regions: entorhinal, inferior temporal, middle temporal, inferior parietal, fusiform, and precuneus. We also included the volume of one visual region, lateral occipitalcortex, under the hypothesis that visual impairment may cause atrophy in visual tracts and brain regions.(38,39) MRI was obtained from the first imaging visit (n=36,855), which was acquired using identical scanners (Siemens Skyra 3T scanner with a standard 32-channel head coil) and the same protocol across sites.(40) Volumetric, cortical thickness, and surface area measures were obtained using FreeSurfer. All volumetric measures were adjusted for intracranial volume.

### Genotyping and genetic risk scores for cataracts, myopia, and AD

Samples were genotyped in batches using 2 closely related arrays (UK BiLEVE array and UK Biobank Axiom array), as described elsewhere.(41,42) We selected genetic instruments for MR analyses and calculated genetic risk score based on previously published genome-wide association studies (GWAS) for cataracts(18), myopia (19) and AD(17). A multiethnic meta-analysis of genome-wide association studies (GWAS) of cataracts (18) identified genome-wide significant loci in cohorts from UK Biobank and the Genetic Epidemiology Research in Adult Health and Aging (GERA). A genetic risk score for cataracts (Cataracts-GRS) used in primary analyses was calculated based 43 genome-wide significant SNPs (p < 1e-08) from a 23andMe replication study, and independent of each other by removing SNPs in linkage disequilibrium (r^2^>0.3)(**supplemental table 2**). We calculated the Cataracts-GRS by multiplying each individual’s risk allele count for each locus by the effect estimates for that SNP from the GWAS (347,209 cases and 2,887,246 controls) and summing scores for all 43 SNPs to create a weighted sum.(43) We evaluated a secondary genetic risk score for cataracts from the study’s smaller GERA subcohort of participants with European ancestry (28,092 cases and 50,487 controls), which was completely independent from SNPs identified in UKB to minimize any weak instrument bias.(44) This GRS was limited to 7 SNPs, after restricting to SNPs that were significant (p< 1e-07) and removing SNPs in linkage disequilibrium. Both of these final genetic risk scores were transformed into a Z-score for interpretability.

The same approach was used to create a GRS for myopia from a meta-analysis identifying loci associated with myopia in UK Biobank and GERA cohorts.(19) The genetic risk score for myopia (Myopia-GRS) used in primary analyses was limited to 55 SNPs that replicated in 23andMe and were statistically significant (p < 1e-08 from the 23andMe replication study, and independent of each other by removing SNPs in linkage disequilibrium [r^2^>0.3]), and it used effect estimates obtained from the 23andMe replication study (**supplemental table 3**). Secondary analyses used a Myopia-GRS (106 SNPs) created from the study’s GERA subcohort (p < 1e-08 and removed SNPs in linkage disequilibrium [r^2^>0.3]).

Finally, a GRS for AD (AD-GRS) was created using effect estimates from a large GWAS on dementia that did not include the UKB.(17) We focused on genetic risk for AD to represent the reverse causation between incipient dementia and vision since genetic risk for AD is well-described and AD comprises the largest subtype of dementia. The final AD-GRS was a weighted sum of 27 SNPs, including two apolipoprotein E (*APOE*) ε4 alleles (**supplemental table 4**). We replicated our analyses using a version of the AD-GRS without the *APOE* alleles.

The genome-wide association studies used for cataracts, myopia, and AD were all adjusted for age, sex, and main principal components. Our genetic analyses adjust for the same covariates. Additionally, we confirmed that each GRS (Cataracts-GRS, Myopia-GRS, AD-GRS) was associated with its corresponding exposure in our UKB dataset (**supplemental table 5**), an important criteria for valid MRs.(45)

### Covariates

Age at visit (baseline or imaging), self-reported sex (male, female), self-reported racial and ethnic identity (Asian, Black, Chinese, Mixed, Other, White), quintiles of index of multiple deprivation by country of origin (highest, above average, average, below average, and lowest), and 6 indicators for history of relevant comorbidities (falls, broken bones, cardiovascular disease, stroke, diabetes, and problems hearing) were reported at baseline assessment. Genetic models (including GRS and MR models) were adjusted for age at visit, sex, and the first 10 genetic ancestry principal components to adjust for confounding by population stratification. Imaging models included adjustment for imaging center.

### Statistical Analysis

#### Observational Analyses

We conducted observational analyses to investigate associations between visual acuity, visual conditions, dementia, and cognition within our sample. First, we assessed the relationship between eye problems (cataracts, myopia, binary 20/40 vision, glaucoma, diabetic retinopathy, and age-related macular degeneration) and all-cause dementia, AD, and VaD using using Cox proportional-hazard models censored for death, loss to follow-up, or end of study period. In a secondary model focused on cataracts, we evaluated whether history of cataract surgery modified associations with dementia risk. Second, we assessed whether the cataracts, myopia, cataracts-GRS, and myopia-GRS were associated with cross-sectional differences in cognitive assessments (‘Snap’ test and Pairs Matching) using linear regressions.

Finally, we investigated potential biological pathways for the association between eye problems and dementia by looking at cross-sectional differences in imaging markers using linear regressions adjusted for confounders and imaging center. Confounders included in models were demographics and health comorbidities. Models including visual acuity and myopia were additionally adjusted for glasses use which could correct visual impairment.

#### Mendelian Randomization

To better investigate causality compared to the observational covariate adjustment, we used genetic risk to evaluate the relationships between cataracts, myopia, and dementia with observed outcomes. There are four main assumptions that need to be met for MR validity.(45) First, the relevance condition requires that the genetic variants are associated with the risk factor or exposure (for example, SNPs in the AD-GRS must be associated with AD). Second, the independence assumption requires that there are no unmeasured confounders of the SNPs and the outcome. Third, the exclusion restriction assumption states that the only effect of the SNPs on the outcome operates through the exposure. Finally, we must assume monotonicity, meaning that our defined GRS does not have opposite effects on individuals within our sample. This limits the generalizability of the MR effect to “compliers.”(46)

We first modeled the relationships of two vision GRS (Cataracts-GRS and Myopia-GRS) with all-cause dementia using logistic regressions. To evaluate reverse causation, we additionally modeled the relationship between AD-GRS and three vision outcomes (cataracts, myopia, and binary 20/40 vision) using logistic regressions. The use of these GRS allows us to leverage individual level outcome data to examine potential relationships. Next, to further examine significant findings identified by GRS, we used two-sample MR estimators and conducted sensitivity analyses to evaluate MR assumptions. Using the SNPs from each GRS described above as instruments we conducted 2-sample MR assessing the following five directional relationships: 1) from cataracts to all-cause dementia, 2) cataracts to VaD, 3) AD to cataracts, 4) AD to myopia, and 4) AD to 20/40 vision. SNP-outcome associations were estimated in UK Biobank using the same covariate adjustments as the external GWAS for exposure data and reference alleles were harmonized. We report estimates from four MR approaches:(20) MR Egger, weighted median, inverse variance weighted, and weighted mode. Additionally, we report the MR-Egger intercept and the MR-PRESSO global test as tests for horizontal pleiotropy. There were too few SNPs and low precision to use the secondary Cataracts-GRS from GERA in two-sample MR analyses. All analyses were conducted in R (version 1.4.1717), with the MR analysis utilizing the TwoSampleMR package (version 0.5.6).

## RESULTS

Our full analytic sample included 304,953 participants (mean [SD] age at baseline, 62.06 [4.08]; 54.5% female). Of this sample, over an average follow-up of 12.71 years, 7,155 incident dementia cases were identified (7,676 when adding additional diagnoses from EHR). When restricting to individuals with available data, 14,295 (4.7%) participants reported a history of cataracts, 19,670 had myopia (28.2%), and 2,754 had a binary visual acuity 20/40 vision or worse (3.9%)(**table 1**).

**Table 1.** Characteristics of UK Biobank participants included in analyses.

| **Participant Characteristics** | **N with observed data** | **Mean (SD) or n (%)** |
| --- | --- | --- |
| Age (years) | 304,953 | 62.06 (4.08) |
| Female | 304,953 | 163,825 (53.72) |
| Primary Cataracts-GRS | 150,858 | 0.03 (0.98) |
| Myopia-GRS | 150,858 | -0.02 (1) |
| AD-GRS with *APOE* | 183,439 | 0.01 (1) |
| Self-reported cataracts | 304,953 | 14,295 (4.69) |
| Self-reported cataract surgery | 304,953 | 2,335 (0.77) |
| Self-reported glaucoma | 304,953 | 6,187 (2.03) |
| Self-reported AMD | 304,953 | 3,321 (1.09) |
| Self-reported diabetic retinopathy | 304,953 | 2,850 (0.93) |
| Myopia | 69,852 | 19,670 (28.16) |
| Worse than 20/40 vision | 71,429 | 2,754 (3.86) |
| Snap Reaction Time (ms) | 301,362 | 580.88 (120.04) |
| Number of Incorrect Pairs Matches | 302,402 | 2.52 (1.95) |
| Total brain volume (mm<sup>3</sup>) | 36,591 | 1,152,218.9 (109,824.54) |
| Lateral occipital volume (mm<sup>3</sup>) | 36,855 | 25,261.47 (3,347.28) |
| Hippocampal volume (mm<sup>3</sup>) | 36,855 | 7,998.23 (791.8) |

Abbreviations: GRS = genetic risk score; AD = Alzheimer disease; AMD = age-related macular degeneration

### Vision impairments and eye conditions associated with cognition and risk of dementia

Glaucoma and AMD were not associated with all-cause dementia, though confidence intervals were wide and consistent with moderate increased risk. Diabetic retinopathy was strongly associated with increased hazard of all-cause dementia (HR= 1.63, 95% CI: 1.40-1.91), AD (HR= 1.41, 95% CI: 1.00-1.97, and VaD (HR= 1.77, 95% CI: 1.30-2.41; **supplemental table 6**). Results did not differ meaningfully when excluding additional EHR diagnoses of dementia.

Binary 20/40 visual acuity was associated with increased hazard of all-cause dementia (HR=1.35, 95% CI: 1.06-1.70), but not AD or VaD. Additionally, myopia was associated with decreased hazard of dementia (HR = 0.92, 95% CI: 0.81-1.05; **table 2**), although this was not statistically significant.

**Table 2.** Cox models for the associations between eye conditions and all-cause dementia, AD, and VaD (including additional EHR diagnoses). Models include adjustment for: age at visit, self-reported sex, self-reported racial and ethnic identity, index of multiple deprivation by country of origin, and binary indicators for history of falls, broken bones, cardiovascular disease, stroke, diabetes, and problems hearing.

| **Eye condition** | **n** | **All-Cause Dementia HR (95% CI)** | **AD HR (95% CI)** | **VaD HR (95% CI)** |
| --- | --- | --- | --- | --- |
| Cataracts | 280,527 | 1.18<br>(1.07-1.29) | 1.25<br>(1.07-1.47) | 1.25<br>(1.01-1.55) |
| Myopia* | 64,003 | 0.92<br>(0.81-1.05) | 1.06<br>(0.85-1.34) | 0.74<br>(0.50-1.11) |
| Binary 20/40 Vision* | 65,429 | 1.35<br>(1.06-1.70) | 1.18<br>(0.75-1.85) | 1.57<br>(0.85-2.90) |

\*Additionally adjusted for glasses use.

Cataracts were associated with all-cause dementia (HR = 1.18, 95% CI: 1.07-1.29), AD (HR=1.25, 95% CI: 1.07-1.47), and VaD (HR=1.25, 95% CI: 1.01-1.55; **table 2**). When additionally adjusting for cataract surgery (n=2,581, 0.5%), cataracts were still associated with increased odds of all-cause dementia (HR = 1.19, 95% CI: 1.08-1.32). Cataract surgery was not associated with dementia (OR = 0.99, 95% CI: 0.57-1.70) compared to not having cataract surgery, although confidence intervals for this interval were quite wide. There was no statistically significant interaction between cataracts and cataract surgery (p=0.72).

The primary cataracts-GRS was not significantly associated with cognitive scores, but increased genetic risk for myopia had a small but statistically significant association with better visual memory (**supplemental table 7**).

### Associations with brain MRI volumes

Cataracts were associated with smaller total grey matter volume (β: −2483.27 mm^3^, 95% CI: −4225.21, −741.34; **fig 2, supplemental table 8**), smaller lateral occipital volume (β: −243.47 mm^3^, −416.81, −70.12), and greater white matter hyperintensity volume (β: 531.00 mm^3^, 95% CI: 79.87, 982.13). Cataracts-GRS was significantly associated with smaller total grey matter volume (β: −375.17 mm^3^, 95% CI: −680.10, −70.24) and total brain volume (β: - 597.43 mm^3^, 95% CI: −1077.87, −117.00). There was no evidence that cataracts or genetic risk for cataracts were associated with brain regions linked to AD-related cortical thinning (**supplemental table 7**).

**Fig 2.**
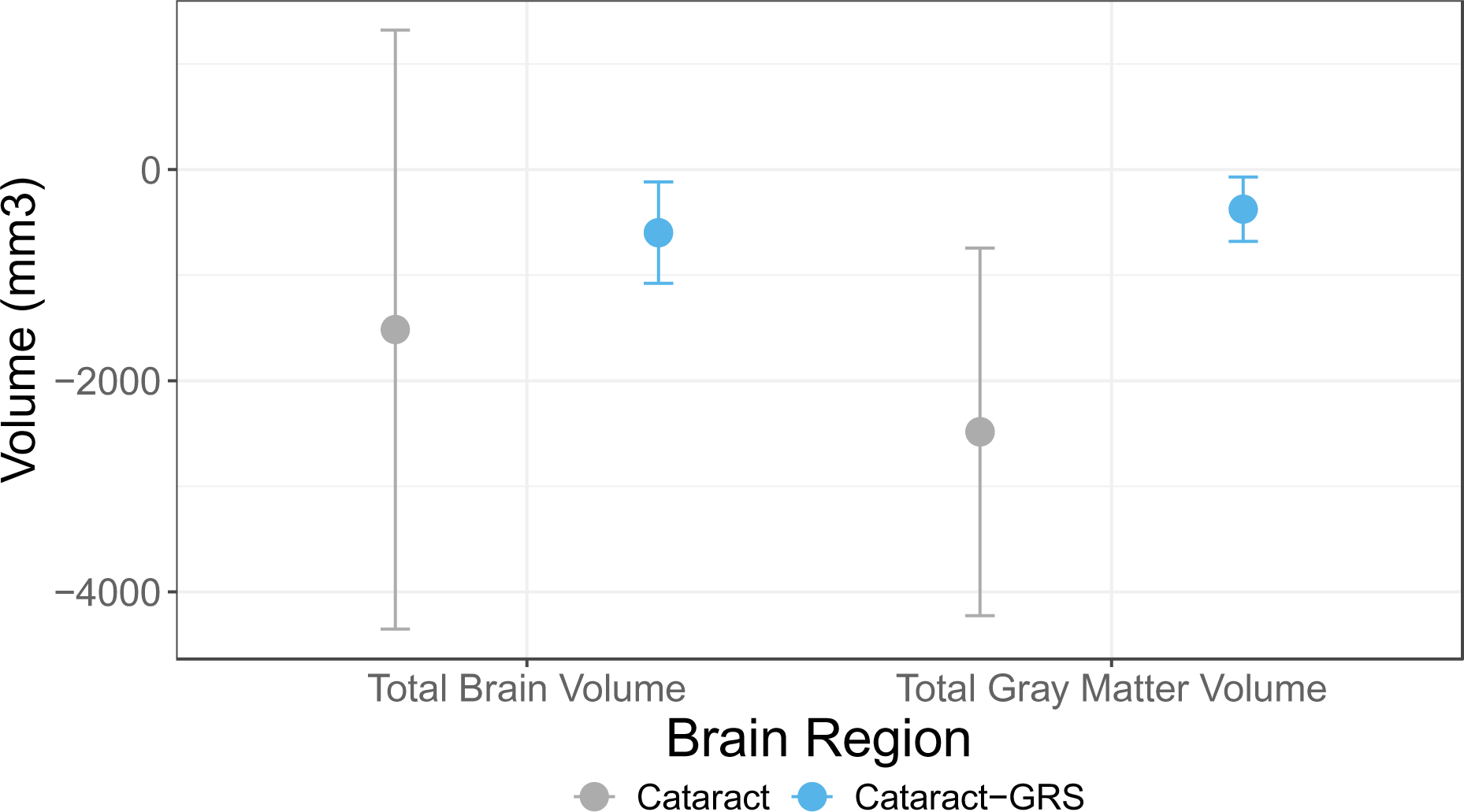
Association between cataracts or cataracts-GRS and total brain and gray matter volumes (mm^3^). Models for cataracts include adjustment for: age at visit, self-reported sex, self-reported racial and ethnic identity, index of multiple deprivation by country of origin, binary indicators for history of comorbidities (falls, broken bones, cardiovascular disease, stroke, diabetes, and problems hearing), and imaging center. Models for cataracts-GRS (primary) include adjustment for: age, sex, first 10 principal components, imaging center, and intracranial volume.

### Mendelian Randomization

Genetic risk for cataracts (primary Cataracts-GRS) was not associated with increased risk of all-cause dementia in UK Biobank (OR = 1.03, 95% CI: 0.99-1.06). When examining dementia subtypes, cataracts-GRS was associated with VaD (OR = 1.10, 95% CI: 1.04-1.16), but not AD (OR = 1.01, 95% CI: 1.04-1.16; **supplemental table 9**). In secondary analyses using GRS from GERA (secondary cataract-GRS), cataracts-GRS was strongly associated with all-cause dementia (OR=1.23, 95% CI: 1.20-1.26), AD (OR=1.30, 95% CI: 1.25-1.35), and VaD (OR = 1.20, 95% CI: 1.14-1.26; **supplemental table 9**). The Myopia-GRS (primary and secondary) was not associated with all-cause dementia, AD, or VaD.

When evaluating the reverse direction, AD-GRS was associated with lower odds of myopia (OR = 0.97, 95% CI: 0.95-0.99), but not with cataracts (OR = 0.99, 95% CI: 0.97-1.01). When removing APOE, AD-GRS was no longer associated with myopia (OR=0.98, 95% CI: 0.96-1.01), but it was associated with small decreases in odds of self-reported and ICD diagnoses of cataracts (OR=0.98, 95% CI: 0.97-0.99; **supplemental table 10**).

Using MR (on our primary cataract-GRS), estimates for the causal relationship of cataracts on dementia were all greater than 1 (OR estimates ranging 1.21-1.37; **table 3**), although none of these estimates were statistically significant. Additionally, there was no evidence of horizontal pleiotropy using the MR-Egger method (β = −0.01, p=0.586). The effect of individual cataracts SNPs on all-cause dementia is shown in **Fig 3**, with no evidence for large outliers. MR estimates indicate a stronger causal effect of cataract on VaD (OR estimates ranging 1.92-2.33; **table 3**), and all estimates were statistically significant except for MR-Egger (OR = 2.33, 95% CI: 0.90-6.02). There was additionally no evidence of horizontal pleiotropy (MR-Egger intercept = −0.01, p=0.658). Results did not change when including additional EHR diagnoses of dementia in the outcome. MR-PRESSO provided similar estimates (OR = 1.21, 95% CI: 0.98-1.50; Global test for horizontal pleiotropy p= 0.37).(47)

**Figure 3.**
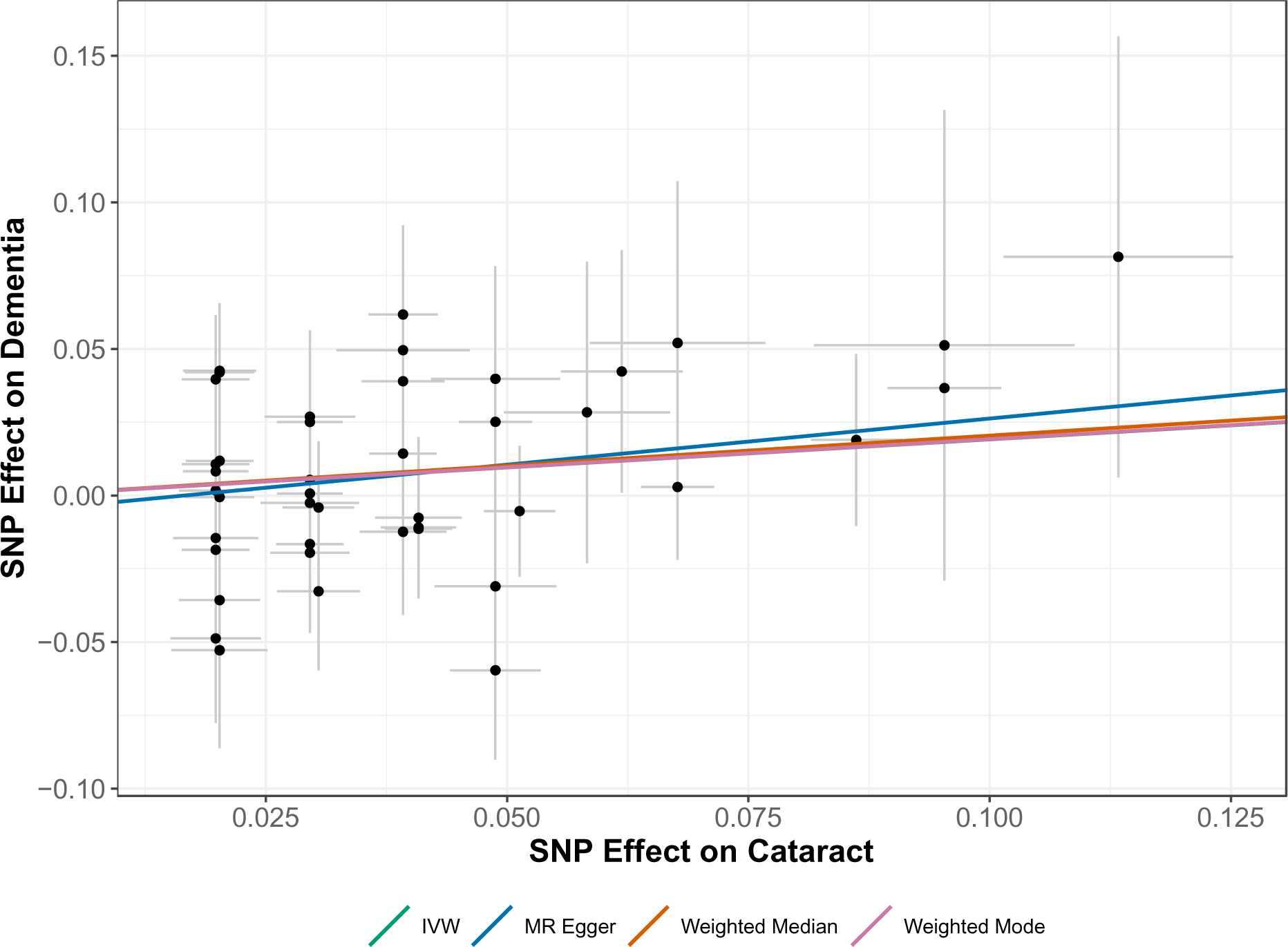
**Effect of 43 cataracts SNPs on both cataracts and dementia**. MR estimates for the effect of cataracts (primary) are shown for inverse variance weighted (IVW), MR-Egger (which adjusts for pleiotropy), weighted-median, and weighted-mode.

**Table 3.**
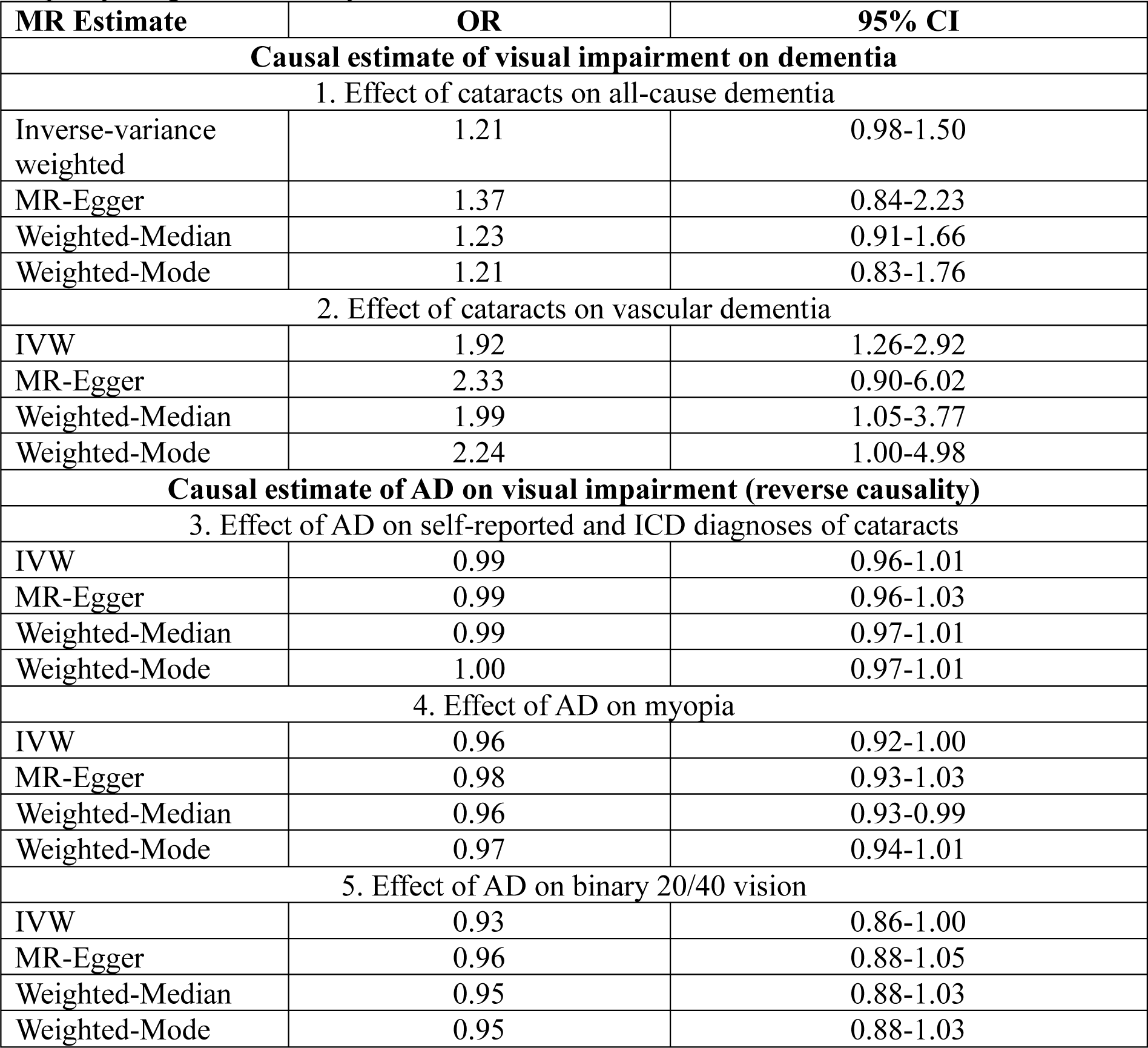
Summary MR estimates. . The effect of cataracts on (1) algorithmic dementia and (2) algorithmic VaD with additional diagnoses, (3) the effect of AD on self-reported and ICD diagnoses of cataracts, (4) the effect of AD on myopia, and (5) the effect of AD on binary 20/40 vision. Models adjusted for age, sex, and first 10 principal components to proxy for genetic ancestry.

To evaluate whether dementia also causes later visual changes, we also checked for reverse causation using MR. AD SNPs were not significantly associated with cataracts, myopia, or binary 20/40 vision (**table 3**) across the majority of summary MR estimates, which trended towards the inverse direction. There was no evidence of horizontal pleiotropy for the MR between AD and cataracts (β = −0.003, p=0.392)(**supplemental fig 1**).

## DISCUSSION

We evaluated the relationships between vision impairments and dementia risk in the large UK Biobank using observational and MR approaches. A history of cataracts and poor visual acuity were strongly associated with dementia risk. In our MR studies, higher genetic risk for cataracts was associated with increased risk of VaD, and our secondary cataract genetic risk score was associated with all-cause dementia and AD. Using 2-sample MR estimators, cataracts increased VaD risk 2-fold. Genetic risk for AD was not associated with later visual impairment or eye conditions, suggesting observational associations are not due to reverse causation. We found evidence that cataracts may be associated with brain atrophy, specifically in metrics reflecting global grey and white matter health. Finally, myopia was associated with decreased risk of dementia, but we found no evidence that this relationship is causal, suggesting that associations between poor visual acuity and dementia are not driven by propensity for myopia. Together these results are generally consistent with the hypothesis that cataracts increase risk of dementia.

Our results extend current evidence by demonstrating that the relationship between cataracts and dementia may be causal and that cataracts are associated with total brain volumes. Consistent with our work several observational studies have found that the presence of cataracts were associated with increased dementia risk,(4,5,48,49). Some previous work found no association between cataracts and dementia(50,51), but was underpowered. One prior MR study did not find a strong causal link between cataracts and AD (22), but did not examine other dementia outcomes. Our findings suggest that associations may be strongest with VaD, as point estimates from MR were about two times larger in magnitude compared to all-cause dementia.

Our findings are strengthened by evaluating multiple ADRD-related outcomes including neuroimaging makers. Few prior studies have examined associations between vision and ADRD-related neurodegeneration (38,39,52). In our study, cataracts were associated with increased brain atrophy, including total grey matter and occipital volumes. While cataracts may lead to broad neurodegeneration, we did not find evidence for AD-related neurodegeneration. As history of cataracts were associated with VaD and white matter hyperintensity volumes (a marker of small vessel ischemic disease),(53) there may be vascular mechanisms linking cataracts and dementia. However, future studies will need to clarify the underlying mechanisms.

Our results are largely consistent with previous literature showing that poor visual acuity(6–8) is associated with increased dementia risk. However, visual acuity was not associated with any brain MRI measures, in contrast to another study.(38) Myopia was associated with decreased risk of dementia in observational models. Although this is consistent with some prior work,(54) we believe this is likely due to residual confounding, as the Myopia-GRS was not associated with all-cause dementia, AD, or VaD. Similar to other studies we also found diabetic retinopathy (55,56)was associated with dementia risk. The causal relationship between these other eye conditions and ADRD should be investigated in future work.

The validity of our MR analyses depends on certain assumptions of IV.(20,21,45) First, we confirmed genetic instruments predict each exposure of interest. Second, while we can’t prove the independence assumption is met, we have adjusted for the strongest potential confounder, principal components as measures of genetic ancestry.(57) Third, we also believe the exclusion restriction assumption holds in all of our MR models, as there was no evidence for horizontal pleiotropy using the MR-Egger Intercept. Finally, the monotonicity assumption limits the generalizability of the effect estimates we’ve obtained and may not represent the true magnitude of the effect of cataracts on dementia in different populations.

This study has some important limitations. First, like all analytic models, MR assumptions cannot be made with certainty. Additionally, they are traditionally underpowered, which is reflected in the wide confidence intervals in some of our MR estimates. However, by also including observational methods and multiple GRS, this study still suggests there is a causal effect of cataracts on dementia by triangulation.(58) Second, the genetic analyses included in this study were restricted to individuals in UK Biobank with European ancestry, meaning genetic results may not be generalizable to individuals from other genetic backgrounds. Finally, the individuals in UK Biobank are relatively young (mean age of 62 years) and healthy,(59) leading to only a smaller number of cases of ADRD at this point in the cohort’s follow-up. This study should be replicated in an older cohort with more diverse representation. Our study also has important strengths compared to prior work, including the strength of causal inference through MR, a large sample, and an investigation into potential biological pathways.

In conclusion, this study is an important addition to the literature of eye conditions, visual impairment, and later dementia risk. We found that cataracts but not myopia were associated with increased risk of dementia in both observational and MR analyses. We were able to analyze bidirectional relationships and demonstrated that genetic propensity for dementia likely does not cause later cataracts. These results support the conclusion that treating or preventing cataracts may reduce dementia risk. Future studies should investigate the specific mechanisms, such as vascular disease, that connect cataracts to dementia.

## STUDY FUNDING

This work was supported by National Institutes of Health/National Institute on Aging grants T32AG049663-06A1 (ELF, MT) and K01AG062722 (WDB). The content is solely the responsibility of the authors and does not necessarily represent the official views of the National Institutes of Health. SJA is supported by K99/R00 AG070109l. KY is supported by R35 AG071916. MMG is supported by R01AG057869. HC is supported by the National Eye Institute (NEI) grant R01 EY033010.

## ETHICS APPROVAL

Ethics approval for data collection was obtained from the National Health Service National Research Ethics Service, and all participants provided written informed consent. This research was conducted under UK Biobank project #78748.

## DATA AVAILABILITY STATEMENT

Researchers can apply to access the data used in this study from UK Biobank (http://www.ukbiobank.ac.uk/).

## ACKNOWLEDGEMENTS

We thank the UK Biobank participants and staff. The UK Biobank project is funded by the Medical Research Council, The Wellcome Trust, the Department of Health for England and Wales, the North West Regional Development Agency, and the Scottish Executive. Preliminary sections of this manuscript were presented as poster abstracts at the Society for Epidemiologic Research in June 2023 and the Alzheimer’s Association International Conference in July 2023, and as an oral presentation at the Gerontological Society of America Annual Scientific Meeting in November 2023.

## COMPETING INTERESTS

All authors have completed the ICMJE uniform disclosure form at http://www.icmje.org/disclosure-of-interest/ and declare: support from the above mentioned funders for the submitted work (NIH), no financial relationships with any organizations that might have an interest in the submitted work in the previous three years; no other relationships or activities that could appear to have influenced the submitted work.

## Notes

### Competing Interest Statement

The authors have declared no competing interest.

