## Supplemental Tables and Figures for "Relationships of visual impairment and eye conditions with imaging markers, cognition, and diagnoses of dementia: a bi-directional Mendelian randomization study"

**SUPPLEMENTAL MATERIAL: TABLE OF CONTENTS**

**Supplemental Table 1:** ICD Codes used to extract additional AD and ADRD diagnoses from primary care and in-patient electronic health records through 2021.

**Supplemental Table 2.** SNPs and their log OR effect estimates included in the primary Cataracts-GRS.

**Supplemental Table 3.** SNPs and their log OR effect estimates included in the primary Myopia-GRS.

**Supplemental Table 4.** SNPs and their log OR effect estimates included in the AD-GRS.

**Supplemental Table 5.** Associations between genetic risk scores (Cataracts-GRS, Myopia-GRS, and AD-GRS) with their respective outcomes in UK Biobank.

**Supplemental Table 6.** Cox models for the associations between glaucoma, AMD, and diabetic retinopathy with all-cause dementia, AD, and VaD (including additional EHR diagnoses).

**Supplemental Table 7**. Association between cataracts, cataracts-GRS, myopia, or myopia-GRS and cognitive scores.

**Supplemental Table 8**. Association between cataracts or cataracts-GRS and brain regions (mm^3^) associated with dementia or vision.

**Supplemental Table 9**. Association between primary GRS (obtained from the 23andMe replication study) for cataracts or myopia, and incident dementia, AD, and VaD among participants older than 60 at baseline.

**Supplemental Table 10**. Association between secondary GRS (obtained from GERA cohort) and incident dementia, AD, and VaD among participants older than 60 at baseline.

**Supplemental Table 11**. Associations between AD-GRS, with and without *APOE*, and vision outcomes.

**Supplemental Table 12**. Summary reverse causation MR estimates for: 1) the effect of AD on self-reported and ICD diagnoses of cataracts, 2) the effect of AD on myopia, and 3) the effect of AD on binary 20/40 vision.

**Supplemental Figure 1**. Association between cataracts or cataracts-GRS and total brain and gray matter volumes (mm^3^).

**Supplemental Figure 2**. Effect of 27 AD SNPs on both cataracts and dementia.

**Supplemental Table 1.** ICD Codes used to extract additional AD and ADRD diagnoses from primary care and in-patient electronic health records through 2021.

| **Description** | **ICD-10 code** | **Subtype** |
| --- | --- | --- |
| Dementia in Alzheimer's disease | F00 | AD |
| Dementia in Alzheimer's disease with early onset | F00.0 | AD |
| Dementia in Alzheimer's disease with late onset | F00.1 | AD |
| Dementia in Alzheimer's disease, atypical or mixed type | F00.2 | AD |
| Dementia in Alzheimer's disease, unspecified | F00.9 | AD |
| Alzheimer’s disease | G30 | AD |
| Alzheimer’s disease with early onset | G30.0 | AD |
| Alzheimer’s disease with late onset | G30.1 | AD |
| Other Alzheimer's disease | G30.8 | AD |
| Alzheimer's disease unspecified | G30.9 | AD |
| Vascular dementia | F01 | ADRD |
| Vascular dementia of acute onset | F01.0 | ADRD |
| Multi-infarct dementia | F01.1 | ADRD |
| Subcortical vascular dementia | F01.2 | ADRD |
| Mixed cortical and sub-cortical vascular dementia | F01.3 | ADRD |
| Other vascular dementia | F01.8 | ADRD |
| Vascular dementia, unspecified | F01.9 | ADRD |
| Binswanger's disease | I67.3 | ADRD |
| Dementia in Picks disease | F02.0 | ADRD |
| Circumscribed brain atrophy | G31.0 | ADRD |
| Sporadic Creutzfeldt-Jakob disease | A81.0 | ADRD |
| Dementia in Creutzfeldt-Jacob disease | F02.1 | ADRD |
| Dementia in Huntington’s disease | F02.2 | ADRD |
| Dementia in Parkinson’s disease | F02.3 | ADRD |
| Dementia in HIV disease | F02.4 | ADRD |
| Mental and behavioural disorders due to use of alcohol - amnesic syndrome | F10.6 | ADRD |
| Dementia in other diseases classified elsewhere | F02 | ADRD |
| Dementia in other specified diseases classified elsewhere | F02.8 | ADRD |
| Unspecified dementia | F03 | ADRD |
| Delirium superimposed on dementia | F05.1 | ADRD |
| Senile degeneration of brain | G31.1 | ADRD |
| Other specified degenerative diseases of nervous system | G31.8 | ADRD |

**Supplemental Table 2.** SNPs and their log OR effect estimates included in the primary Cataracts-GRS.

| **Variant** | **Chromosome** | **Locus** | **Effect Allele** | **Effect Estimate** |
| --- | --- | --- | --- | --- |
| rs2073017 | 1 | *CASZ1* | C | -0.0202027 |
| rs3176459 | 1 | *CDKN2C* | G | -0.0202027 |
| rs71646944 | 1 | *ADGRL2* | T | 0.03922071 |
| rs10633030 | 1 | *FAM46C* | CT | 0.0295588 |
| rs2982459 | 1 | *LINC00970* | G | -0.0304592 |
| rs12593 | 1 | *ADCK3* | T | 0.03922071 |
| rs890069 | 2 | *Near TRIB2* | T | -0.0202027 |
| rs10210444 | 2 | *PLB1* | A | 0.03922071 |
| rs7604689 | 2 | *LRP1B-KYNU* | C | -0.0304592 |
| rs62237590 | 3 | *RARB* | C | 0.01980263 |
| rs35256080 | 3 | *ATXN7* | AT | 0.01980263 |
| rs10663094 | 3 | *SOX2-OT* | ACT | 0.04879016 |
| rs72868578 | 4 | *C4orf22-BMP3* | A | 0.06765865 |
| rs7744813 | 6 | *KCNQ5* | A | 0.01980263 |
| rs73015318 | 6 | *QKI* | A | 0.04879016 |
| rs10258092 | 7 | *CREB5* | C | 0.0295588 |
| rs17172647 | 7 | *IGFBP3-TNS3* | G | 0.06765865 |
| rs62621812 | 7 | *ZNF800* | A | 0.11332869 |
| rs12114462 | 8 | *BIN3-EGR3* | C | 0.01980263 |
| rs1679013 | 9 | *CDKN2B-DMRTA1* | T | 0.0295588 |
| rs4742654 | 9 | *FKTN-TAL2* | T | 0.01980263 |
| rs4837205 | 9 | *ST6GALNAC4-PIP5KL1* | C | -0.0202027 |
| rs1014607 | 10 | *BAMBI-LINC01517* | A | -0.040822 |
| rs2274224 | 10 | *PLCE1* | C | 0.0295588 |
| rs73386631 | 11 | *ODF3-BET1L* | T | 0.05826891 |
| rs150648223 | 11 | *5*′ *LOC338694* | ATTT | 0.09531018 |
| rs17739338 | 12 | *CAPRIN2* | T | -0.0618754 |
| rs17608087 | 12 | *MVK-FAM222A* | G | 0.04879016 |
| rs7154613 | 14 | *STXBP6* | T | 0.0295588 |
| rs2855530 | 14 | *BMP4* | C | 0.0295588 |
| rs72714121 | 15 | *OCA2* | T | 0.04879016 |
| rs12901945 | 15 | *RORA-VPS13C* | A | 0.01980263 |
| rs10500355 | 16 | *RBFOX1* | A | 0.03922071 |
| rs73530148 | 16 | *ALDOA* | T | 0.03922071 |
| rs73568154 | 16 | *WWP2* | A | 0.01980263 |
| rs8074331 | 17 | *RHOT1 - RHBDL3* | A | -0.0202027 |
| rs7207025 | 17 | *near MIR2117HG* | A | -0.0202027 |
| rs9038 | 17 | *9-Sep* | C | -0.040822 |
| rs9895741 | 17 | *NPLOC4* | G | -0.0512933 |
| rs75954926 | 17 | *3*′ *METRNL* | G | 0.0295588 |
| rs61744414 | 19 | *CPAMD8* | T | 0.09531018 |
| rs549768142 | 20 | *JAG1* | GAAAAAAAAAAT | -0.040822 |
| rs4814857 | 20 | *SLC24A3* | G | 0.0861777 |

**Supplemental Table 3.** SNPs and their log OR effect estimates included in the primary Myopia-GRS.

| **Variant** | **Chromosome** | **Position** | **Effect Allele** | **Effect Estimate** |
| --- | --- | --- | --- | --- |
| rs2808515 | 1 | 200346324 | G | 0.0707 |
| rs2017760 | 1 | 207455421 | G | -0.1036 |
| rs300751 | 2 | 209063 | C | -0.0774 |
| rs9309272 | 2 | 56095366 | T | 0.127 |
| rs61049169 | 2 | 146888708 | A | -0.0755 |
| rs192716 | 2 | 157376472 | C | -0.0829 |
| rs2573081 | 2 | 178828507 | G | 0.0646 |
| rs1550094 | 2 | 233385396 | A | -0.1004 |
| rs14165 | 3 | 53847408 | G | 0.0779 |
| rs6549043 | 3 | 85632126 | T | 0.0739 |
| rs6767786 | 3 | 141104180 | A | -0.0664 |
| rs74764079 | 4 | 81952637 | A | 0.2958 |
| rs1309551 | 5 | 64288656 | G | -0.0693 |
| rs7730838 | 5 | 71708698 | T | -0.0627 |
| rs7444298 | 5 | 87730027 | G | 0.0846 |
| rs11758482 | 6 | 2438231 | G | 0.0927 |
| rs4145443 | 6 | 22068174 | T | 0.0729 |
| rs2207136 | 6 | 50809720 | C | -0.0689 |
| rs7744813 | 6 | 73643289 | A | 0.1006 |
| rs12193446 | 6 | 129820038 | G | -0.2244 |
| rs6465760 | 7 | 99591418 | A | 0.069 |
| rs12234576 | 7 | 158928375 | A | -0.1062 |
| rs16890057 | 8 | 40726582 | A | -0.0794 |
| rs72621438 | 8 | 60178580 | G | -0.1064 |
| rs9643433 | 8 | 78952371 | A | -0.0708 |
| rs62538956 | 9 | 12679244 | C | 0.1016 |
| rs11145746 | 9 | 71834380 | A | 0.0969 |
| rs7042950 | 9 | 77149837 | G | 0.0919 |
| rs10826198 | 10 | 60274182 | T | 0.0731 |
| rs12778014 | 10 | 94950273 | A | -0.0835 |
| rs807037 | 10 | 102824349 | C | 0.0695 |
| rs56299331 | 10 | 114788436 | T | 0.0811 |
| rs6484385 | 11 | 28668416 | C | 0.0744 |
| rs11602008 | 11 | 40149305 | T | 0.1204 |
| rs11226856 | 11 | 105699048 | G | 0.0918 |
| rs5442 | 12 | 6954864 | A | 0.1439 |
| rs1468993 | 12 | 46155145 | C | 0.0786 |
| rs3138142 | 12 | 56115585 | T | -0.1191 |
| rs11178462 | 12 | 71269789 | A | -0.1055 |
| rs1328371 | 13 | 93912785 | C | -0.066 |
| rs34217772 | 14 | 42273570 | G | 0.0967 |
| rs34935520 | 14 | 61091401 | A | -0.0772 |
| rs35337422 | 14 | 104407243 | C | 0.1109 |
| rs524952 | 15 | 35005886 | A | 0.1713 |
| rs75227249 | 15 | 48763008 | T | -0.1161 |
| rs1007365 | 15 | 79379492 | G | 0.0853 |
| rs8039459 | 15 | 82320426 | G | 0.0755 |
| rs17648524 | 16 | 7459683 | C | 0.0878 |
| rs4635359 | 16 | 80537760 | C | -0.078 |
| rs2908972 | 17 | 11407259 | A | 0.0886 |
| rs62067167 | 17 | 31251711 | T | 0.1156 |
| rs7222840 | 17 | 47280915 | C | 0.0851 |
| rs12603264 | 17 | 54729518 | T | 0.0765 |
| rs4793501 | 17 | 68718734 | T | 0.0996 |
| rs734559 | 18 | 72179579 | A | -0.0811 |

**Supplemental Table 4.** SNPs and their log OR effect estimates included in the AD-GRS.

| **Variant** | **Chromosome** | **Locus** | **Effect Allele** | **Effect Estimate** |
| --- | --- | --- | --- | --- |
| rs4844610 | 1 | *CR1* | A | 0.157 |
| rs6733839 | 2 | *BIN1* | T | 0.182 |
| rs10933431 | 2 | *INPP5D* | G | -0.094 |
| rs9271058 | 6 | *HLA -DRB1* | A | 0.095 |
| rs75932628 | 6 | *TREM2* | T | 0.732 |
| rs9473117 | 6 | *CD2AP* | C | 0.086 |
| rs114812713 | 6 | *OARD1* | C | 0.278 |
| rs12539172 | 7 | *NYAP1* | T | -0.083 |
| rs10808026 | 7 | *EPHA1* | A | -0.105 |
| rs73223431 | 8 | *PTK2B* | T | 0.095 |
| rs9331896 | 8 | *CLU* | C | -0.128 |
| rs7920721 | 10 | *ECHDC3* | G | 0.077 |
| rs3740688 | 11 | *SPI1* | G | -0.083 |
| rs7933202 | 11 | *MS4A2* | C | -0.117 |
| rs3851179 | 11 | *PICALM* | T | -0.128 |
| rs11218343 | 11 | *SORL1* | C | -0.223 |
| rs17125924 | 14 | *FERMT2* | G | 0.131 |
| rs12881735 | 14 | *SLC24A4* | C | -0.083 |
| rs593742 | 15 | *ADAM10* | G | -0.073 |
| rs7185636 | 16 | *IQCK* | C | -0.083 |
| rs62039712 | 16 | *WWOX* | A | 0.148 |
| rs138190086 | 17 | *ACE* | A | 0.262 |
| rs3752246 | 19 | *ABCA7* | G | 0.140 |
| rs429358 | 19 | *APOE* | C | 1.351 |
| rs7412 | 19 | *APOE* | T | -0.386 |
| rs6024870 | 20 | *CASS4* | A | -0.128 |
| rs2830500 | 21 | *ADAMTS1* | A | -0.073 |

**Supplemental Table 5.** Associations between genetic risk scores (Cataracts-GRS, Myopia-GRS, and AD-GRS) with their respective outcomes in UK Biobank. Models include adjustment for age, sex, and first 10 principal components.

| **Genetic Risk Score** | **Outcome** | **n** | **Adjusted OR**  **(95% CI)** |
| --- | --- | --- | --- |
| *Primary Analysis* | | | |
| Primary Cataracts-GRS | Self-Reported Cataracts | 150,858 | 1.18  (1.16-1.21) |
| Primary Cataracts-GRS | Self-Reported and ICD Diagnoses of Cataracts | 150,858 | 1.21  (1.19-1.23) |
| Primary Myopia-GRS | Assessment-Derived Myopia | 34,157 | 1.44  (1.40-1.47 |
| AD-GRS | Incident Algorithmic ADRD | 183,439 | 1.80  (1.76-1.84) |
| *Secondary Analyses* | | | |
| Secondary Cataracts-GRS | Self-Reported Cataracts | 183,439 | 1.05  (1.03-1.07) |
| Secondary Cataracts-GRS | Self-Reported and ICD Diagnoses of Cataracts | 183,439 | 1.05  (1.04-1.06) |
| Secondary Myopia-GRS | Assessment-Derived Myopia | 40,665 | 0.72  (0.71-0.74) |
| AD-GRS without *APOE* | Incident Algorithmic ADRD | 183,439 | 1.20  (1.17-1.24) |

**Supplemental Table 6**. Cox models for the associations between glaucoma, AMD, and diabetic retinopathy with all-cause dementia, AD, and VaD (including additional EHR diagnoses). Models include adjustment for: age at visit, self-reported sex, self-reported racial and ethnic identity, index of multiple deprivation by country of origin, and binary indicators for history of falls, broken bones, cardiovascular disease, stroke, diabetes, and problems hearing.

| **Eye condition** | **n** | **All-Cause Dementia HR (95% CI)** | **AD**  **HR (95% CI)** | **VaD**  **HR (95% CI)** |
| --- | --- | --- | --- | --- |
| Glaucoma | 280,527 | 1.13  (0.98-1.30) | 1.00  (0.76-1.30) | 1.25  (0.91-1.74) |
| Age-related Macular Degeneration | 280,527 | 1.17  (0.97-1.41) | 0.94  (0.65-1.35) | 1.25  (0.80-1.95) |
| Diabetic Retinopathy | 280,527 | 1.63  (1.40-1.91) | 1.41  (1.00-1.97) | 1.77  (1.30-2.41) |

**Supplemental Table 7**. Association between cataracts, cataracts-GRS, myopia, or myopia-GRS and cognitive scores. Models for cataracts or myopia include adjustment for: age at visit, self-reported sex, self-reported racial and ethnic identity, index of multiple deprivation by country of origin, and binary indicators for history of comorbidities (falls, broken bones, cardiovascular disease, stroke, diabetes, and problems hearing). Models for cataracts-GRS or myopia-GRS include adjustment for: age, sex, and first 10 principal components.

| **Exposure** | **n** | **Cognitive Assessment** | |
| --- | --- | --- | --- |
|  |  | Reaction Time:  Seconds (95% CI) | Pairs Matching:  No. Incorrect (95% CI) |
| Self-Reported Cataracts | 451,303 | **5.35 (3.27, 7.44)** | **-0.04 (-0.07, -0.01)** |
| Cataracts-GRS | 213,055 | -0.01 (-0.45, 0.44) | -0.002 (-0.01, 0.006) |
| Myopia* | 103,116 | -0.69 (-2.78,1.41) | **-0.06 (-0.10, -0.03)** |
| Myopia-GRS* | 212,906 | -0.18 (-0.67, 0.31) | **-0.01 (-0.02, -0.002)** |

*Additionally adjusted for glasses use.

**Supplemental Table 8**. Association between cataracts or cataracts-GRS and brain regions (mm^3^) associated with dementia or vision. Models for cataracts include adjustment for: age at visit, self-reported sex, self-reported racial and ethnic identity, index of multiple deprivation by country of origin, binary indicators for history of comorbidities (falls, broken bones, cardiovascular disease, stroke, diabetes, and problems hearing), and imaging center. Models for cataracts-GRS include adjustment for: age, sex, first 10 principal components, and imaging center.

| **Exposure** | **Brain Region** | **Coefficient (mm^3^)** | **95% CI** |
| --- | --- | --- | --- |
| Self-reported Cataracts  (n=33,271) | Total brain volume* | -1515.33 | -4351.56, 1320.91 |
|  | Total grey matter volume* | **-2483.27** | **-4225.21, -741.34** |
|  | Hippocampal volume* | -27.11 | -69.47, 15.25 |
|  | White matter hyperintensity volume* | **531.00** | **79.87, 982.13** |
|  | AD-Signature Region | -0.01 | -0.01, 7e-04 |
|  | Lateral occipital volume* | **-243.47** | **-416.81, -70.12** |
| Cataracts-GRS  (n= 26,051) | Total brain volume* | **-597.432** | **-1077.87, -117.00** |
|  | Total grey matter volume* | **-375.17** | **-680.10, -70.24** |
|  | Hippocampal volume* | -4.28 | -11.90, 3.33 |
|  | White matter hyperintensity volume* | 7.60 | -75.67, 90.88 |
|  | AD-Signature Region | -0.0005 | -0.002, 8e-04 |
|  | Lateral occipital volume* | 26.45 | -6.21, 49.75 |

*Additionally adjusted for intracranial volume.

**Supplemental Table 9**. Association between primary GRS (obtained from the 23andMe replication study) for cataracts or myopia, and incident dementia, AD, and VaD among participants older than 60 at baseline. Models adjusted for age, sex, and first 10 principal components.

| **Exposure** | **Outcome** | **n** | **Algorithmic definition of dementia** | | **Composite with additional EHR diagnoses of dementia** | |
| --- | --- | --- | --- | --- | --- | --- |
|  |  |  | Adjusted OR | 95% CI | Adjusted OR | 95% CI |
| Cataracts-GRS | All-cause dementia | 150,858 | 1.03 | 0.99-1.06 | 1.02 | 0.99-1.05 |
| Cataracts-GRS | AD | 150,858 | 1.01 | 0.97-1.06 | 1.00 | 0.96-1.05 |
| Cataracts-GRS | VaD | 150,858 | **1.10** | **1.03-1.17** | **1.09** | **1.03-1.16** |
| Myopia-GRS* | All-cause dementia | 150,765 | 1.01 | 0.98-1.04 | 1.00 | 0.97-1.03 |
| Myopia-GRS* | AD | 150,765 | 1.02 | 0.97-1.07 | 1.01 | 0.97-1.06 |
| Myopia-GRS* | VaD | 150,765 | 1.02 | 0.96-1.08 | 1.01 | 0.95-1.07 |

*Additionally adjusted for glasses use.

**Supplemental Table 10**. Association between secondary GRS (obtained from GERA cohort) and incident dementia, AD, and VaD among participants older than 60 at baseline. Models adjusted for age, sex, and first 10 principal components.

| **Exposure** | **Outcome** | **n** | **Algorithmic definition of dementia** | | **Composite with additional EHR diagnoses of dementia** | |
| --- | --- | --- | --- | --- | --- | --- |
|  |  |  | Adjusted OR | 95% CI | Adjusted OR | 95% CI |
| Cataracts-GRS | All-cause dementia | 183,439 | **1.24** | **1.20-1.27** | **1.23** | **1.20-1.26** |
| Cataracts-GRS | AD | 183,439 | **1.30** | **1.25-1.35** | **1.30** | **1.25-1.35** |
| Cataracts-GRS | VaD | 183,439 | **1.20** | **1.13-1.26** | **1.20** | **1.14-1.26** |
| Myopia-GRS* | All-cause dementia | 183,320 | 1.00 | 0.97-1.02 | 1.01 | 0.99-1.04 |
| Myopia-GRS* | AD | 183,320 | 1.01 | 0.97-1.06 | 1.03 | 0.99-1.07 |
| Myopia-GRS* | VaD | 183,320 | 0.95 | 0.90-1.01 | 0.96 | 0.91-1.01 |

*Additionally adjusted for glasses use.

**Supplemental Table 11**. Associations between AD-GRS, with and without *APOE*, and vision outcomes. Models adjusted for age, sex, and first 10 principal components.

| **Outcome** | **n** | **Adjusted OR (95% CI)** | **Adjusted OR without *APOE* (95% CI)** |
| --- | --- | --- | --- |
| Self-Reported Cataracts | 183,439 | 1.00  (0.98-1.02) | 0.98  (0.96-1.00) |
| Self-Reported and ICD Diagnoses of Cataracts | 183,439 | 0.99  (0.98-1.00) | **0.98**  **(0.97-0.99)** |
| Myopia | 40,687 | **0.97**  **(0.95-0.99)** | 0.98  (0.96-1.01) |
| Binary 20/40 Vision | 41,646 | 0.97  (0.92-1.02) | 1.00  (0.95-1.05) |

**Supplemental Table 12**. Summary reverse causation MR estimates for: 1) the effect of AD on self-reported and ICD diagnoses of cataracts, 2) the effect of AD on myopia, and 3) the effect of AD on binary 20/40 vision. Models adjusted for age, sex, and first 10 principal components to proxy for genetic ancestry.

| **MR Estimate** | **OR** | **95% CI** |
| --- | --- | --- |
| **Causal estimate of AD on visual impairment (reverse causality)** | | |
| 1. Effect of AD on self-reported and ICD diagnoses of cataracts | | |
| IVW | 0.99 | 0.96-1.01 |
| MR-Egger | 0.99 | 0.96-1.03 |
| Weighted-Median | 0.99 | 0.97-1.01 |
| Weighted-Mode | 1.00 | 0.97-1.01 |
| 2. Effect of AD on myopia | | |
| IVW | 0.96 | 0.92-1.00 |
| MR-Egger | 0.98 | 0.93-1.03 |
| Weighted-Median | 0.96 | 0.93-0.99 |
| Weighted-Mode | 0.97 | 0.94-1.01 |
| 3. Effect of AD on binary 20/40 vision | | |
| IVW | 0.93 | 0.86-1.00 |
| MR-Egger | 0.96 | 0.88-1.05 |
| Weighted-Median | 0.95 | 0.88-1.03 |
| Weighted-Mode | 0.95 | 0.88-1.03 |

**Supplemental Figure 1. Association between cataracts or cataracts-GRS and total brain and gray matter volumes (mm^3^).** Models for cataracts include adjustment for: age at visit, self-reported sex, self-reported racial and ethnic identity, index of multiple deprivation by country of origin, binary indicators for history of comorbidities (falls, broken bones, cardiovascular disease, stroke, diabetes, and problems hearing), and imaging center. Models for cataracts-GRS (primary) include adjustment for: age, sex, first 10 principal components, imaging center, and intracranial volume.

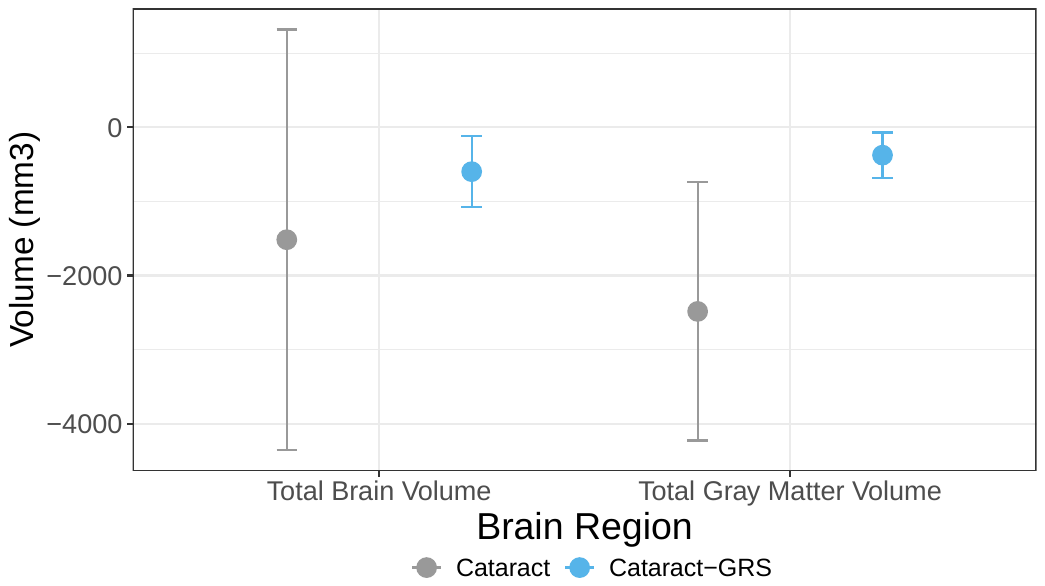

**Supplemental Figure 2**. Effect of 27 AD SNPs on both cataracts and dementia. MR estimates for the effect of AD on cataract are shown for inverse variance weighted (IVW), MR-Egger (which adjusts for pleiotropy), weighted-median, and weighted-mode.

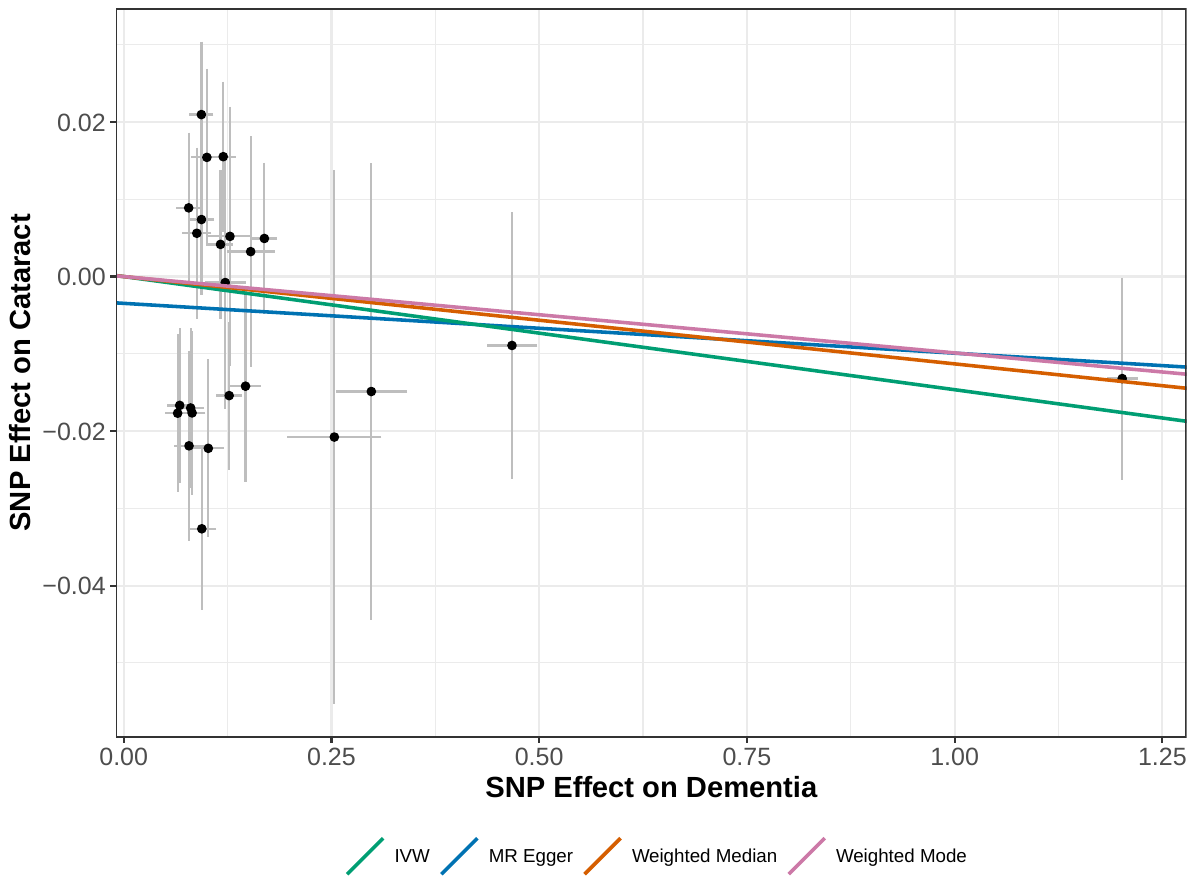
